# Intravesical *Lacticaseicillus rhamnosus* GG reduces symptoms among people with spinal cord injury and disease who use intermittent catheterization: A randomized comparison of two- and four-dose regimens

**DOI:** 10.64898/2026.07.17.26358333

**Authors:** Suzanne L. Groah, Rochelle E. Tractenberg, Christopher R. Riegner, Catherine S. Forster

## Abstract

**Background:** Urinary tract infection (UTI) is the most common secondary condition among people with spinal cord injury/disease (SCI/D). Intravesical *Lacticaseibacillus rhamnosus* GG (LGG) is an antibiotic-sparing approach to managing urinary symptoms.

**Objective:** Determine the optimal number of doses of intravesical LGG for urinary symptom reduction.

**Design:** Prospective, randomized, two-arm dosing trial.

**Setting:** National recruitment with a local subsample providing urine samples in Washington, DC, USA.

**Participants:** Adults with SCI/D and neurogenic lower urinary tract dysfunction (NLUTD) who use intermittent catheterization (IC); 177 enrolled and randomized (intention-to-treat), with 76 compliant instillers (39 low-dose, 37 high-dose) in the per-protocol analytic sample.

**Interventions:** Two (2 doses/24 hours) or four (4 doses/36 hours) intravesical LGG regimens, self-initiated in response to cloudier or malodorous urine per the Self-Management Protocol using Probiotics (SMP-Pro).

**Main Outcome Measures:** Primary: proportion achieving ≥20% reduction on the Urinary Symptom Questionnaire for Neurogenic Bladder–Intermittent Catheter version (USQNB-IC). Secondary: urinary biomarkers (leukocyte esterase, nitrite, white blood cells, urinary neutrophil gelatinase-associated lipocalin [uNGAL]) and standard urine culture (SUC) in a local subsample.

**Results:** By Day 2, 57.9% (63.8% low-dose; 51.2% high-dose) achieved ≥20% total symptom reduction; high-dose success rose to 70.0% by Day 4. Thirty percent of high-dose participants did not respond at either time point and could not be distinguished from responders by demographics or urine biomarkers. Urinary biomarkers and SUC were unchanged pre- to post-instillation. No serious adverse events were adjudicated as attributable to intravesical LGG by an independent Data Safety Monitoring Board (DSMB).

**Conclusions:** A two-dose course of intravesical LGG yields clinically meaningful symptom improvement in the majority of people with SCI/D and NLUTD who use IC; four doses benefits a meaningful subgroup of two-day non-responders, while a small cohort remains nonresponsive. These results provide preliminary dosing guidance and support progression to a definitive trial.

## Introduction

Urinary tract infection (UTI) is the most common outpatient infection world-wide; for people with spinal cord injury or disease (SCI/D) and neurogenic lower urinary tract dysfunction (NLUTD), it is the most common infection (≈2.5/person/year)^1^, secondary condition, cause for emergency-room visits, and infectious cause of hospitalization.^2–5^ The standard of care is reactive: diagnostic delays drive repeated antibiotic exposures, overtreatment of presumed UTI, and poor antibiotic stewardship—all of which fuel the prevalence of multi-drug resistant organisms (MDROs) in this population.^6^ The high burden of MDROs among people with SCI/D^6^—associated with longer hospitalizations,^4^ increased mortality,^7^ higher costs,^8^ and reduced functional gains^9^—mirrors the world-wide antimicrobial resistance crisis^8,10–12^ reflecting the harms of indiscriminate antibiotic use under diagnostic uncertainty.^13^

The growing recognition that the urinary tract harbors a resident microbiome (urobiome)—and that disruption of this urobiome may predispose to infection—has prompted our study of live biotherapeutic approaches that manage early urinary symptoms proactively, before they progress to UTI requiring antibiotics.^14,15^ We have advanced the use of intravesically instilled *Lacticaseibacillus rhamnosus* GG (LGG; formerly *Lactobacillus rhamnosus* GG)^16^ as an approach to avoiding UTI by treating the most common urinary symptoms that are bothersome to the patient (cloudier, more foul-smelling urine)^17,18^ but do not rise to the level of UTI according to the Infectious Diseases Society of America.^19^ This approach is antibiotic-sparing and proactive, and intravesical LGG can be self-administered by individuals with SCI/D. It stems from our initial discovery that *Lactobacillus* is a predominant genus in the urobiome of some healthy controls,^20^ while it is absent from the urobiomes of many people with SCI/D in favor of a predominance of potential uropathogens such as *Escherichia*, *Klebsiella*, and *Enterococcus*.^20,21^

In our first-in-human trial, intravesical LGG was safe and well-tolerated,^15,22,23^ with preliminary efficacy showing an average within-person reduction in total symptom burden of at least 10%.^24^ Building on those findings, the present study estimated and compared the proportions of participants achieving clinically meaningful symptom reduction (≥20%) across two dose levels of intravesical LGG among people with SCI/D and NLUTD who manage their bladders with intermittent catheterization (IC).

## Methods

The trial was approved by the institutional review board. The protocol conformed to the 1975 Declaration of Helsinki, and intravesical LGG was administered under FDA Investigational New Drug (IND) #16306. An independent Data Safety Monitoring Board (DSMB) reviewed adverse events at six-month intervals.

This prospective, randomized (1:1) dosing trial compared the effect of 2 doses (in 24 hours) versus 4 doses (in 36 hours) of intravesical LGG on urinary symptoms. Our primary outcome was a binary variable representing the yes/no achievement of clinically meaningful change in total urinary symptom burden. Symptoms were assessed using the Urinary Symptoms Questionnaire for Neurogenic Bladder for individuals using intermittent catheterization (USQNB-IC),^25^ one of a set of urinary symptoms assessment instruments developed for, and validated with, individuals with NLUTD due to SCI/D.^17,18,25^ We defined “success” as ≥20% reduction in USQNB-IC total symptom burden, which was estimated every 24 hours after treatment initiation. Secondary outcomes were the effects of LGG dose on standard urinary markers (urine WBC, nitrite, leukocyte esterase), urinary neutrophil gelatinase-associated lipocalin (uNGAL), and cultivable bacteria. Initiation of the intervention was triggered by cloudier or more malodorous urine than typical, as guided by the SMP-Pro.^15^

A total of 1,450 individuals were screened. The CONSORT diagram appears in Figure 1.

**Fig. 1.**
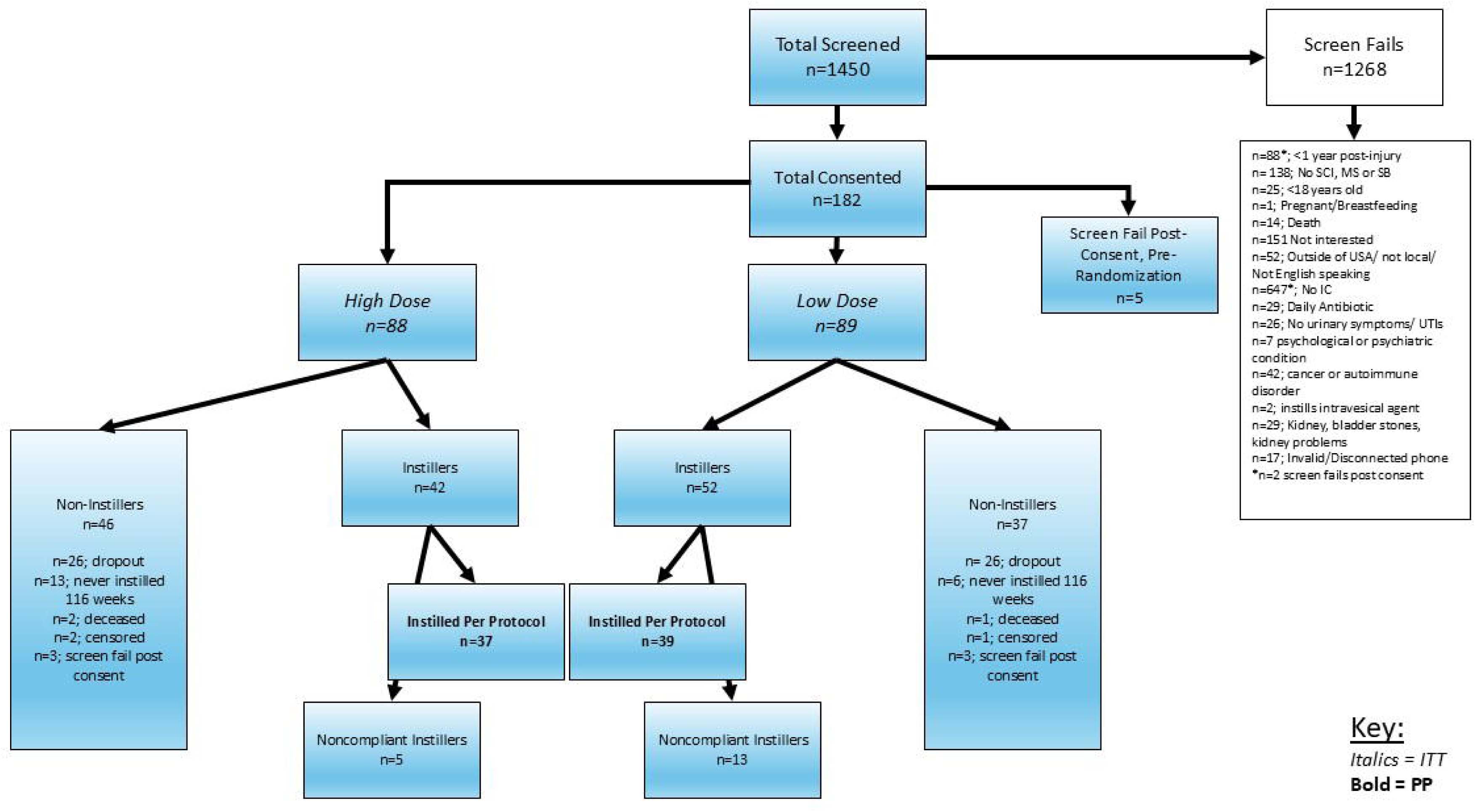
CONSORT.

Participants were recruited nationally and were eligible if they: (1) were ≥18 years of age; (2) had SCI/D for at least 1 year; (3) had NLUTD; (4) used IC for bladder management; (5) had a history of ≥2 episodes of urinary symptoms in the past year; and (6) were community-dwelling. Exclusion criteria were: (1) known genitourinary pathology beyond NLUTD (e.g., vesicoureteral reflux, bladder or kidney stones); (2) use of prophylactic antibiotics; (3) instillation of intravesical agents to reduce UTI; (4) immunodeficiency; (5) any antibiotics within the past 2 weeks; (6) psychologic or psychiatric conditions influencing the ability to follow instructions; and (7) participation in a confounding study.

After informed consent, participants were randomized by the biostatistician to low (2 doses in 24 hours) or high (4 doses in 36 hours) dose intravesical LGG using a permuted-blocks procedure generated *a priori*. Each participant received a training manual and attended a 1-on-1 training session on the SMP-Pro and intravesical LGG instillation with a person with lived experience. Participants were provided intervention materials (sterile gloves, new catheters, LGG capsules, 0.9% normal saline, sterile cups, 60-cc syringes, and bed pads). Participants remained in the study for up to 29 months or until SMP-Pro trigger symptoms occurred, at which point pre-instillation symptoms were recorded, LGG was instilled per the randomized regimen, and post-instillation urine symptoms were recorded within 24 hours of the last dose of LGG. Participants did not repeat the intervention. A subset of participants local to the study center also contributed urine for biomarker and culture analyses (secondary outcomes) pre-instillation and post-instillation at the same time as symptoms were recorded. A four-item patient satisfaction survey developed during our prior work^15^ was also administered after the post-instillation urine sample for those who initiated instillation.

Local participants contributed a 50–100-mL urine sample collected under sterile conditions from a new, unused intermittent catheter for uNGAL, WBC, nitrite, and cultivable bacterial species (in colony-forming units, CFU). Standard urine culture (SUC) was performed by standard laboratory technique, which includes inoculation onto MacConkey and blood agar and aerobic incubation at 35–37°C for ≥24 hours. The SUC outcome was defined as growth of any bacteria ≥10^5^ CFU/mL. Urine for uNGAL was centrifuged, aliquoted, and frozen at –80°C within 8 hours of collection; uNGAL was measured by ELISA (Bioporto, Grusbakken, Denmark) in duplicate at a designated reference laboratory.^26,27^

### Statistical analysis

The primary goal of this dose-finding study was to compare the proportion of participants achieving treatment success---defined a priori as a ≥20% reduction in USQNB-IC total symptom burden---between the 2-dose (low) and 4-dose (high) arms. The target sample size was based on this primary comparison. Assuming a two-sided α = 0.05 and 80% power, 54 evaluable participants per arm were required to detect the anticipated between-arm difference in success proportions (30% in the low-dose vs 56.5% in the high-dose arm); this sample also provided 80% power to detect an odds ratio as small as 4.2 in a model adjusting for a single covariate such as sex. To accommodate an anticipated 40% rate of attrition and non-use—participants who remained symptom-free, and therefore did not instill, during the 29-month observation period—the enrollment target was inflated to 91 participants per arm (182 total). We pre-specified a simple proportions test to compare success at 2 versus 4 doses and explored: (a) individuals who responded within 2 doses; (b) those in the high-dose group who did not respond after 2 doses but did after 4; and (c) whether response could be predicted from urinary markers (in the local subsample) or demographics. Since the two dose groups could only be compared at the completion of their allocated doses, we did not correct for multiple inferences.

Three analytic populations were pre-specified: (i) the enrolled and randomized cohort (N=177); (ii) the per-protocol (PP) sample of compliant instillers (n=76), defined as participants who endorsed at least one SMP-Pro trigger symptom and instilled LGG according to the randomized regimen with paired pre- and post-instillation symptom data; and (iii) all others, including those who, according to the SMP-Pro (i.e., per protocol) never instilled. Main analyses focused on the ITT population (compliant and non-compliant instillers); sensitivity analyses for the main outcome featured imputing "non responder" for ITT participants without final data (ITT_imp_), and analyzing the PP sample to determine if conclusions based on PP deviated from those based on the ITT sample. Urine-specific analyses focused on the PP sample. All analyses were performed in SPSS (IBM, 2024).

## Results

### Participant flow and baseline characteristics

Of the 1,450 individuals screened, 177 were enrolled and randomized (Figure 1). Nineteen participants completed the 29-month observation period without experiencing SMP-Pro trigger symptoms and therefore never instilled LGG. There were 76 per protocol instillers, 18 non-compliant instillers and 83 non-instillers (of whom 19 never instilled over the course of the study, 52 withdrew, 3 were censored and 3 died before instilling). While 18 participants failed to comply with the protocol, 13 of these completed the post-instillation symptom survey, and were included in the ITT analyses. Baseline demographic and clinical characteristics by dose group and instillation/study status are presented in Table 1. The two dose groups were similar with respect to age, race/ethnicity, diagnosis, and injury level, although the low-dose group had a higher proportion of males (63.5% vs 50.0%). The local subsample contributing paired urine samples for biomarker and SUC analyses was distributed evenly between dose groups (20 low-dose, 18 high-dose). Seventy-six participants who experienced trigger symptoms and complied with the instillation protocol constitute the PP analytic sample (39 low-dose and 37 high-dose). Their demographic profiles were not significantly different than for the full sample (data not shown).

**Table 1.**
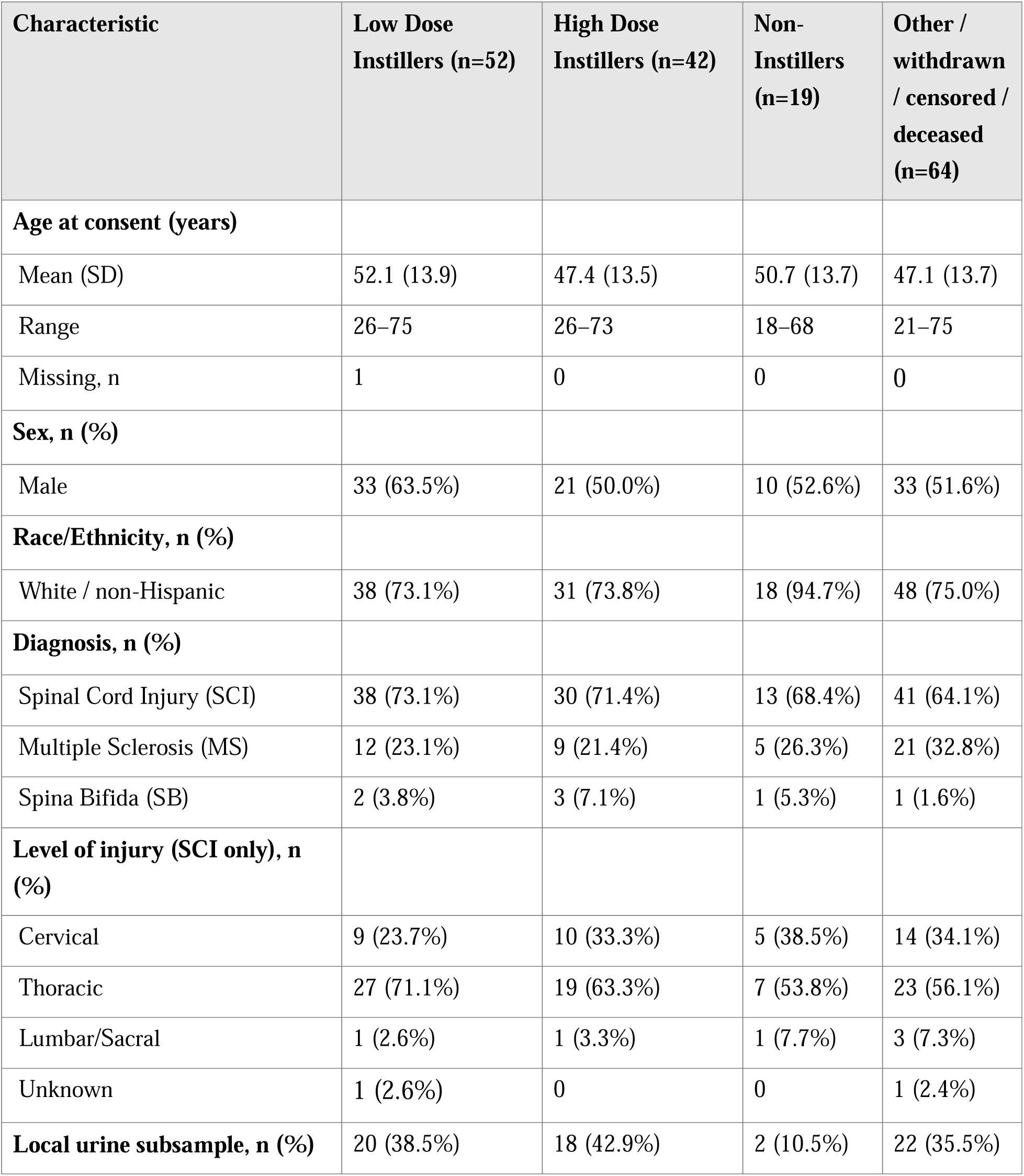
Demographic and clinical characteristics by dose group and study status (ITT/enrolled cohort, N=177).

### Safety and tolerability

All adverse events were systematically reviewed by an independent DSMB at six-month intervals throughout the study. None of the 15 serious adverse events (SAE) were adjudicated by the DSMB as attributable to intravesical LGG, and no protocol-defined stopping criteria were triggered. Three deaths occurred among randomized participants during the observation period; all occurred prior to any instillations; and all were adjudicated by the DSMB as unrelated to the study intervention and consistent with the medical complexity expected in a community-dwelling SCI/D cohort. Among the 177 ITT participants, 8 of the 15 SAEs were determined to be UTIs. Of these 8 UTI SAEs, six occurred prior to any instillation. The two UTI-related hospitalizations occurring among instillers occurred within 30 days of instillation, one each per dosage group. The low dose instiller was noncompliant with the protocol and instilled despite the presence of actionable symptoms (which directed participants not to instill and to consider seeking medical attention), while the high dose instiller ceased the protocol after two instillations (representative of compliance with protocol). Standard urine culture results both grew ≥10^5^ CFU/mL of *E. coli*.

### Effect of intravesical LGG on symptoms

Our *a priori* threshold for success was at least a 20% reduction in symptoms post-instillation, relative to the symptoms at the time of initial instillation. Because there were only two trigger symptoms, change in trigger symptoms could be reported only in discrete values (0%, ±50%, or ±100%). Table 2 shows the proportions achieving the success threshold by dose group and time point in the ITT, per protocol (PP), and imputed non-responder analyses (ITTimp).

**Table 2.** Response rates by analysis population.

| Timepoint / Group | Criterion | ITT |  | PP |  | ITTimp |  |
| --- | --- | --- | --- | --- | --- | --- | --- |
|  |  | n<sup>†</sup> | % | n | % | n | % |
| Day 2 — Low dose | Total | 30/47 | 63.8% | 22/39 | 56.4% | 30/52 | 57.7% |
|  | Trigger* | 22/47 | 46.8% | 18/39 | 46.2% | 22/52 | 42.3% |
| Day 2 — High dose | Total | 21/41 | 51.2% | 19/36 | 52.8% | 21/42 | 50.0% |
|  | Trigger* | 12/40 | 30.0% | 12/36 | 33.3% | 12/42 | 28.6% |
| Day 2 — Combined | Total | 51/88 | 58.0% | 41/75 | 54.7% | 51/94 | 54.3% |
|  | Trigger* | 34/87 | 39.1% | 30/75 | 40.0% | 34/94 | 36.2% |
| Day 4 — High dose | Total | 28/40 | 70.0% | 25/36 | 69.4% | 28/42 | 66.7% |
|  | Trigger* | 24/39 | 61.5% | 24/36 | 66.7% | 24/42 | 57.1% |

ITT = intention-to-treat (observed cases); PP = per-protocol; ITTimp = ITT with "non-responder" imputed for missing post-instillation outcomes. Success defined a priori as $\geq 20\%$ reduction in symptoms after instillation.
<sup>†</sup>Some participants missing 2nd symptom survey; correct n in denominator. \* Trigger-symptom change reported in discrete values (0%, $\pm 50\%$ , $\pm 100\%$ ) given two trigger items.

Forty individuals in the high-dose ITT group had USQNB-IC data at day 4 and of these, 12 (30%) did not achieve the 20% reduction at day 2 or day 4 (one high-dose participant met success at day 2 but by day 4 was no longer ≥20% lower than pre-instillation). Among non-responders with final-day USQNB-IC data in either dose group, no demographic signal predicted non-response (sex, diagnosis, injury level; all p>0.05 before correction for multiple comparisons).

Among ITT low-dose responders at day 2 (30/47, 63.8%), the within-person reduction averaged 43.3% in USQNB-IC urine symptoms, 9.2% in bladder symptoms, 6.1% in actionable symptoms, and 6.7% in constitutional symptoms. Among ITT high-dose responders at day 2 (21/41, 51.2%), the within-person reduction averaged 47.0% in urine symptoms, 4.6% in bladder symptoms, 1.5% in actionable symptoms, and 7.2% in constitutional symptoms. There were no statistically significant differences (all p>0.05).

### Effect of intravesical LGG on urinary biomarkers and bacterial growth

Local participants contributed paired urine samples (pre- and post-instillation) for nitrite (NIT, positive/negative), leukocyte esterase (LE; negative; trace; 1+, 2+, 3+), and urine WBC (none through packed, by HPF). For the clearest perspective on the effects of the intervention, we focus these analyses on compliant participants (PP). Both responders and non-responders showed high rates of positive nitrite, elevated LE, and pyuria at the time of trigger symptoms (Table 3). The most common outcome in all four cells (responder/non-responder × low/high dose) was no change. ITT analyses showed the same patterns (data not shown).

**Table 3.**
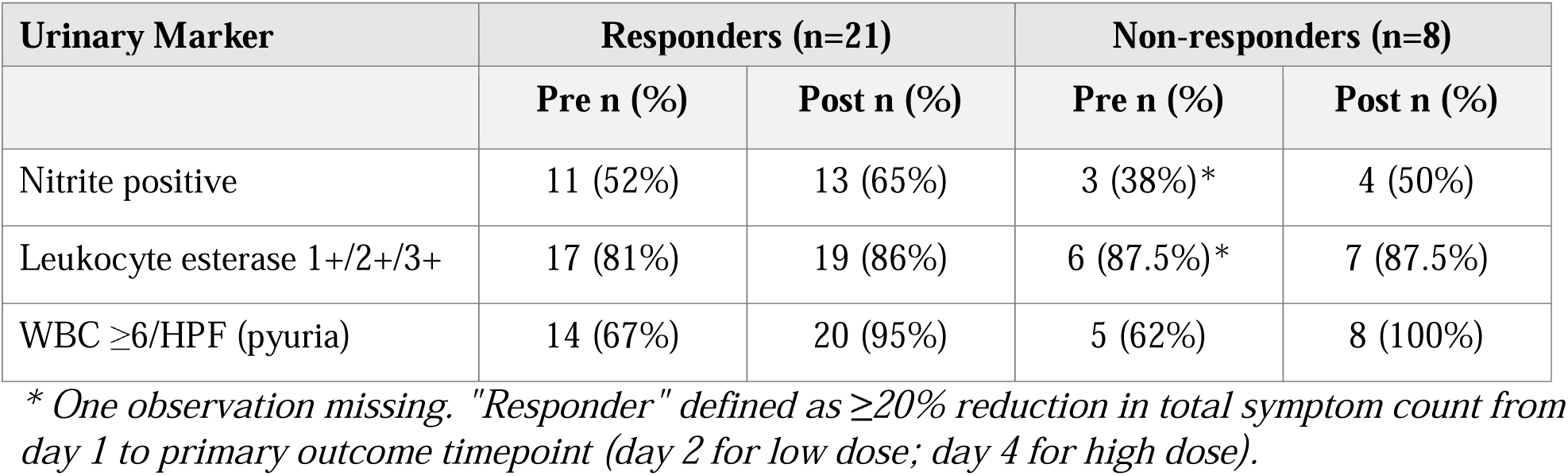
Pre- and post-instillation urinary markers among responders and non-responders in the PP local subsample (collapsed across dose groups).

There were no differences between dose groups in the number of organisms cultured at ≥10^5^ CFU/mL pre- or post-instillation; the majority of cultures (10/14 low-dose, 12/15 high-dose) yielded one organism at this level both before and after instillation. The most commonly cultured organism (>10^5^ CFU/mL) in both groups, pre- and post-instillation, was *E. coli*. Other common uropathogens (*K. pneumoniae*, *E. faecalis*, *P. aeruginosa*) were present pre-instillation but were less likely to be detected in the high-dose group post-instillation. *Lactobacillus* was cultured in one individual in the low-dose group and in two individuals in the high-dose group (*L. rhamnosus*/*Lactobacillus* sp.) post-instillation (Table 4).

**Table 4.** Cultivable organisms isolated at ≥10 CFU/mL before and after intravesical *Lactobacillus rhamnosus* GG (LGG) instillation, by dose group (local paired-urine subsample).

| Organism | Low Pre<br>(n=14) | Low Post<br>(n=14) | Low $\Delta$ | High Pre<br>(n=15) | High Post<br>(n=15) | High $\Delta$ |
| --- | --- | --- | --- | --- | --- | --- |
| ***Gram-negative potential uropathogens*** |  |  |  |  |  |  |
| *E. coli* | 8 | 7 | -1 | 7 | 9 | +2 |
| *K. pneumoniae* | 2 | 2 | 0 | 2 | 0 | -2 |
| *E. cloacae* | 1 | 1 | 0 | 1 | 0 | -1 |
| *A. baumannii* (complex) | 1 | 1 | 0 | 1 | 0 | -1 |
| *P. aeruginosa* | 1 | 0 | -1 | 0 | 0 | 0 |
| *A. urinae* | 0 | 0 | 0 | 0 | 1 | +1 |
| ***Gram-positive potential uropathogens*** |  |  |  |  |  |  |
| *E. faecalis* | 0 | 1 | +1 | 1 | 0 | -1 |
| ***Probiotic / intervention organisms*** |  |  |  |  |  |  |
| *L. rhamnosus* | 0 | 0 | 0 | 0 | 1 | +1 |
| *Lactobacillus* sp. | 0 | 0 | 0 | 0 | 1 | +1 |
| ***Commensal / skin or oral flora*** |  |  |  |  |  |  |
| *S. mitis* | 0 | 1 | +1 | 1 | 0 | -1 |
| *S. α-hemolytic* | 0 | 0 | 0 | 0 | 1 | +1 |
| Unidentified isolate | 0 | 0 | 0 | 0 | 1 | +1 |

In the PP sample, pre-instillation uNGAL ranged from 9.79 to 495.1 ng/mL (mean 144.4, SD 140.8) and post-instillation values ranged from 10.9 to 1,478.0 ng/mL (mean 212.8, SD 371.3); the mean within-person change was +71.9 ng/mL (SD 352.8, range -353.4 - 1276.1 ng/mL), with substantial within-person variability. ITT descriptive analyses were essentially identical to the PP uNGAL distributions pre, post, and change, confirming that compliance status did not influence uNGAL and that uNGAL did not move directionally with treatment in responders or non-responders of either dose group (Figure 2). Given small sex-stratified sample sizes, inferential testing was not performed.

**Figure 2.**
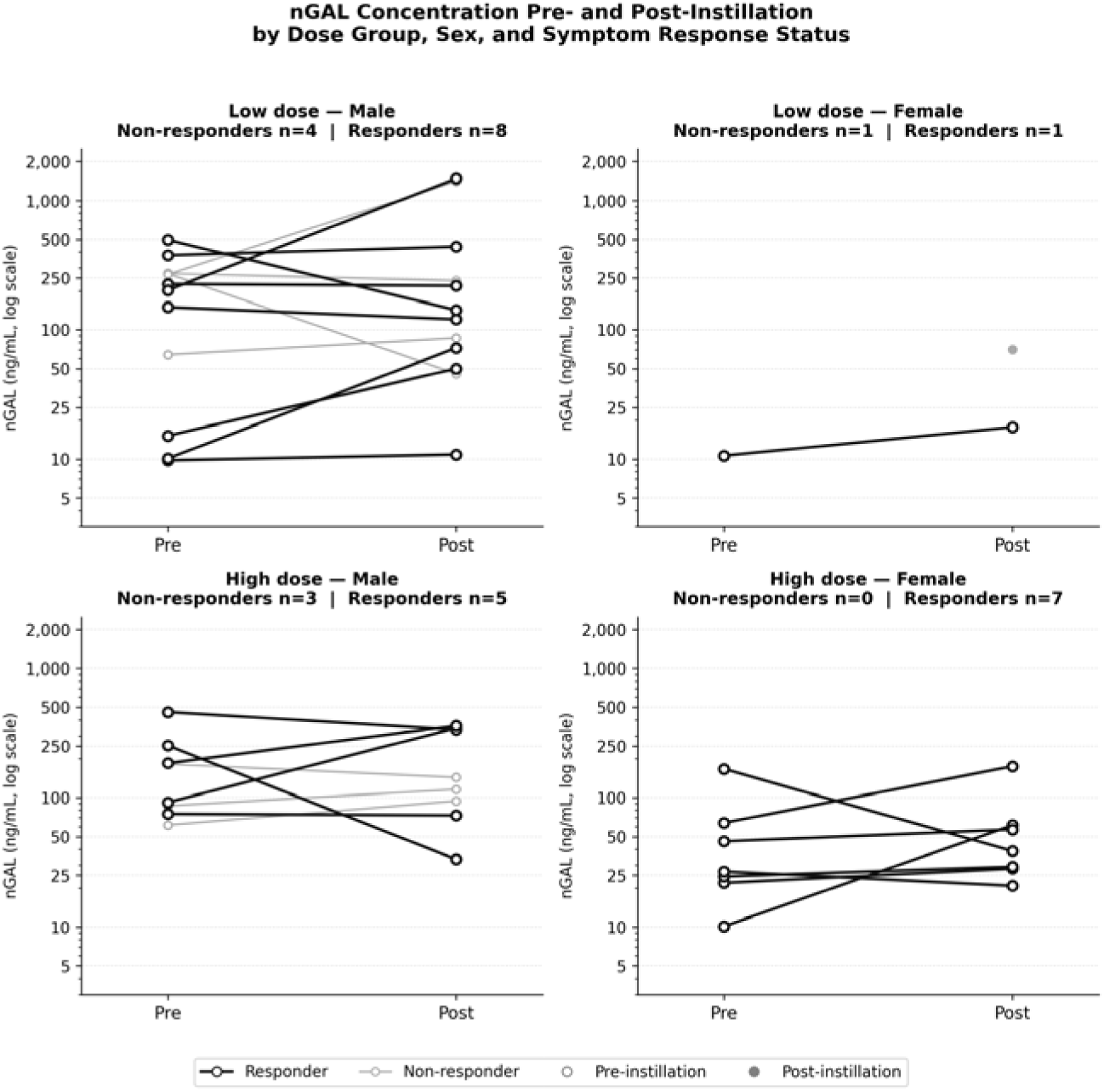
Urinary nGAL values (log scale) for PP responders and non-responders by dose group and sex, before the first instillation and 24 hours after the last instillation.

All participants who instilled, whether or not they were compliant with the SMP-Pro, were administered a four-item survey 24 hours after their last dose. Three items asked for ratings on a 5-point Likert scale of perceived change in 1) frequency; 2) severity; and 3) impact of the intervention on their symptoms. Collapsing across dose groups but focusing on individuals who followed the protocol, the modal response of women (n=36) was satisfied or very satisfied with the effects of intravesical LGG on symptom frequency (44.4%), severity (58.3%), and impact (47.2%). Men were similarly satisfied or very satisfied with the effects of the intervention on frequency (51.9%), severity (61.5%), and impact (57.7%) of their symptoms. Among women, 19.4%-38.9% reported no detectible change on these three symptom dimensions; among men, 25.0%-38.5% reported no detectible change. The remaining responses were unsatisfied/very unsatisfied, and these were the smallest proportions of men and women on all three dimensions. These did not vary based on whether the participant was a responder (achieved 20% or more reduction in symptoms) or not. We also asked participants to give a likelihood rating from 0 to 100 that indicated their willingness to seek out and pay for the intervention. Collapsing across sex and dose group, the average rating given by those who had achieved the 20% success criterion by day 2 (n=51) was 60.12 (SD=31.26), while for those who had not achieved this criterion by day 2 (n=37) the average rating was 59.27 (SD=32.2). At day 4, the average rating from those achieving success (n=36) was 66.64 (SD=28.62) while for those who did not achieve the 20% reduction in total symptoms by day 4 (n=11), the average rating was 55.92 (SD=34.81). No inference tests were planned or done on these ratings.

## Discussion

In this randomized study of intravesical LGG among adults with NLUTD due to SCI/D who use IC, a two-dose regimen produced clinically meaningful total symptom reduction (≥20%) in approximately 58% of all instillers within 24 hours of completing the second of two instillations. For those who continued with a four-dose regimen, the response rate increased to 70%, with nine individuals (ITT) meeting the success criterion after 2 additional doses. When considering the effects on the two trigger symptoms (cloudier/fouler smelling urine), 47% and 62% of ITT participants in the low- and high-dose arms, respectively, achieved at least 50% reduction (because there were only two options for improvement for two symptoms, 50% and 100%). The patterns of "success" were similar for men and women and irrespective of number of doses. Thirty percent of participants in the four-dose group did not respond at either time point, and these non-responders could not be distinguished from responders on the basis of demographics, etiology of SCI/D, or any urinary markers. Thus, a short course of two doses suffices for the majority of individuals, while four doses offers additional benefit in a meaningful subgroup.

Several converging lines of evidence support a biologically plausible mechanism for the symptom relief observed following intravesical LGG. *Lactobacillus* species compete directly with uropathogens for urothelial adhesion,^28,29^ produce antimicrobial substances (lactic acid, hydrogen peroxide, bacteriocins, phenyl-lactic acid) that reduce local pathogen burden without selecting for antibiotic resistance,^29,30^ and inhibit virulence and biofilm formation by uropathogenic *E. coli*^29^—a particularly relevant effect for those who use indwelling catheters, a population who may also benefit from this intervention. Beyond direct antimicrobial activity, *L. rhamnosus* modulates the urothelial innate immune response by potentiating NF-κB activation through TLR4 upregulation^31^ while simultaneously downregulating pro-inflammatory cytokines.^32^ Our prior urobiome analysis demonstrated that one to two intravesical LGG instillations significantly reduced alpha diversity and decreased the relative abundance of *Escherichia/Shigella* and *Aerococcus*, consistent with active ecological remodeling of the bladder microenvironment.^33^ Although the present study did not directly measure these mechanisms, the rapid symptom improvement observed within 24 hours of instillation is consistent with this constellation of effects.

In the present trial, standard urine biomarkers (leukocyte esterase, nitrite, WBC) and uNGAL did not distinguish responders from non-responders or track symptom change.^13,34,35^ This is consistent with the well-described dissociation between urinary biomarkers and symptoms among people with NLUTD due to SCI/D.^13,20,21,36^ uNGAL, in particular, showed wide, sex-dependent spreads with both increases and decreases observed among responders and non-responders, rather than a consistent directional change^36^. Cultures showed modest dose-dependent shifts — lack of or reduced growth of *K. pneumoniae* and other Gram-negative uropathogens in the high-dose post-instillation samples, with concurrent recovery of *L. rhamnosus* and *Lactobacillus* sp. — but these are descriptive observations in a small subsample and were not the subject of inferential testing. These findings reinforce that conventional urinalysis and culture-based markers are insensitive indicators of therapeutic response to LGG in this population, and they underscore the value of validated patient-reported outcome measures—such as the USQNB-IC—as the primary endpoint, alongside continued development of mechanism-informed biomarkers for responder identification.

### Future directions

Longer-term post-dose follow-up of urinary symptoms, the urobiome, and impact on recurrent UTI are high-priority next steps. Central questions for that work are durability of the instilled organism and whether clinical benefit outlasts detectable persistence of the instilled organism. Even though an instilled live biotherapeutic may decline to non-detectable levels over time, durable clinical benefit may persist via beneficial remodeling of the resident urobiome. A close parallel comes from work with intravaginal *Lactobacillus crispatus* live biotherapeutics for recurrent bacterial vaginosis,^37^ in which strain-level analyses suggest that, as the administered strain becomes undetectable, native *L. crispatus* strains expand and persist, providing prolonged colonization resistance against pathogens. Whether an analogous urinary "engraftment-then-replacement" trajectory follows intravesical LGG—and whether maintenance or cyclic re-dosing is required to sustain benefit—are open questions for future investigation. Combined with development of mechanism-informed biomarkers for responder identification, such studies will help define the role of intravesical live biotherapeutics in NLUTD.

### Limitations

Several limitations of this study should be noted. First, this was a randomized trial powered to detect a clinically meaningful difference in success proportions between dose arms. The n=76 compliant instillers analyzed in the per-protocol sample is modestly below the original target, reflecting the 40% non-use/attrition rate anticipated in the protocol and planned for in the original enrollment. Accordingly, the observed difference in response rates between arms should be interpreted with caution and requires confirmation in a larger trial. Second, urinary biomarker data (uNGAL, leukocyte esterase, nitrite, WBC) and SUC were obtained from a local convenience subsample (n=29), limiting the representativeness and inferential utility of those analyses; the biomarker subsample was not powered for hypothesis testing. Third, the study sample was predominantly White and non-Hispanic, which may limit generalizability to the broader and more demographically diverse SCI/D population. Fourth, each participant was eligible for only a single treatment episode; the present design therefore cannot address cumulative, repeated-use, or maintenance effects of intravesical LGG over time, motivating the longer-term follow-up and re-dosing studies described above. Finally, the absence of a non-instilling control arm limits attribution of the observed within-person symptom changes solely to LGG; an ongoing comparative-effectiveness trial of intravesical LGG versus saline bladder wash will provide that comparison.^14^

### Conclusion

Intravesical LGG produces clinically meaningful urinary symptom reduction in the majority of people with SCI/D and NLUTD who use IC, using a validated patient-reported outcome measure as the primary endpoint. Two doses are sufficient for most individuals; extending treatment to four doses increases the cumulative response rate to 70% and should be offered to those who have not responded by day 2. Roughly 30% who complete a four-dose course will not respond—a finding that underscores the need for biomarker-based responder identification in future work. These results provide actionable, evidence-based guidance for intravesical LGG dosing and support progression of this antibiotic-sparing therapeutic approach toward a definitive trial.

## Trial registration and regulatory

ClinicalTrials.gov identifier: NCT04373512

FDA Investigational New Drug (IND) application: #16306

IRB approval: MedStar Health Research Institute IRB #00001124

## Author contributions (CRediT)

SG contributed conceptualization, methodology, resources and led writing of the original draft, review and editing, visualization, project administration and funding acquisition.

RET contributed conceptualization; methodology; software; validation; formal analysis; investigation; writing — original draft; writing — review & editing; visualization; and funding acquisition

CR contributed validation, investigation, data curation, writing – review & editing, visualization, and project administration.

CF contributed to methodology, investigation, and reviewed and edited the manuscript

The authors also acknowledge the contributions of the Data Safety Monitoring Board members.

## Data availability statement

De-identified individual participant data will be available upon reasonable request from the corresponding author following publication, subject to institutional and CDMRP data-sharing requirements.

## Use of artificial-intelligence assistance

Drafting and copy-editing of this manuscript were supported by an AI writing assistant (Anthropic Claude). All scientific content, claims, and interpretations were authored, verified, and approved by the listed authors, who take full responsibility for the integrity of the work.

## References

1. Cardenas DD, Hoffman JM, Kirshblum S, McKinley W. Etiology and incidence of rehospitalization after traumatic spinal cord injury: a multicenter analysis. Arch Phys Med Rehabil. 2004;85(11):11.

2. Cardenas DD, Moore KN, Dannels-McClure A, et al. Intermittent catheterization with a hydrophilic-coated catheter delays urinary tract infections in acute spinal cord injury: a prospective, randomized, multicenter trial. PM R. 2011;3(5):5. doi:10.1016/j.pmrj.2011.01.001

3. Haisma JA, van der Woude LH, Stam HJ, et al. Complications following spinal cord injury: Occurrence and risk factors in a longitudinal study during and after inpatient rehabilitation. J Rehabil Med. 2007;39:393–398. doi:10.2340/16501977-0067

4. Consortium for Spinal Cord Medicine. Bladder management for adults with spinal cord injury: a clinical practice guideline for health-care providers. J Spinal Cord Med. 2006;29(5):5.

5. DeJong G, Tian W, Hsieh CH, et al. Rehospitalization in the First Year of Traumatic Spinal Cord Injury After Discharge From Medical Rehabilitation. Arch Phys Med Rehabil. 2013;94(4):S87–S97. doi:10.1016/j.apmr.2012.10.037

6. Lautenbach E, Patel JB, Bilker WB, Edelstein PH, Fishman NO. Extended-Spectrum - Lactamase-Producing Escherichia coli and Klebsiella pneumoniae: Risk Factors for Infection and Impact of Resistance on Outcomes. Clin Infect Dis. 2001;32(8):8. doi:10.1086/319757

7. García Leoni ME, Esclarín De Ruz A. Management of urinary tract infection in patients with spinal cord injuries. Clin Microbiol Infect. 2003;9(8):8. doi:10.1046/j.1469-0691.2003.00643.x

8. Waites KB, Chen Y, DeVivo MJ, Canupp KC, Moser SA. Antimicrobial resistance in gram-negative bacteria isolated from the urinary tract in community-residing persons with spinal cord injury. Arch Phys Med Rehabil. 2000;81(6):6. doi:10.1016/s0003-9993(00)90108-4

9. Stampas A, Dominick E, Zhu L. Evaluation of functional outcomes in traumatic spinal cord injury with rehabilitation-acquired urinary tract infections: A retrospective study. J Spinal Cord Med. 2019;42(5):5. doi:10.1080/10790268.2018.1452389

10. Antimicrobial resistance. August 28, 2023. Accessed August 28, 2023. https://www.who.int/news-room/fact-sheets/detail/antimicrobial-resistance

11. CDC. Antibiotic Resistance Threatens Everyone. Centers for Disease Control and Prevention. July 20, 2020. Accessed March 19, 2021. https://www.cdc.gov/drugresistance/index.html

12. Dinh A, Saliba M, Saadeh D, et al. Blood stream infections due to multidrug-resistant organisms among spinal cord-injured patients, epidemiology over 16 years and associated risks: a comparative study. Spinal Cord. 2016;54(9):9. doi:10.1038/sc.2015.234

13. Brubaker L, Chai TC, Horsley H, Khasriya R, Moreland RB, Wolfe AJ. Tarnished gold—the “standard” urine culture: reassessing the characteristics of a criterion standard for detecting urinary microbes. Front Urol. 2023;3. Accessed July 31, 2023. https://www.frontiersin.org/articles/10.3389/fruro.2023.1206046

14. Groah SL, Tractenberg RE. Intravesical Lactobacillus rhamnosus GG versus Saline Bladder Wash: Protocol for a Randomized, Controlled, Comparative Effectiveness Clinical Trial. Top Spinal Cord Inj Rehabil. 2022;28(4):4. doi:10.46292/sci22-00005

15. Groah S, Ljungberg I, Tractenberg R, Rounds A. Self-Management of Urinary Symptoms Using a Probiotic in People with Spinal Cord Injuries, Spina Bifida, and Multiple Sclerosis. Patient-Centered Outcomes Research Institute (PCORI); 2020. doi:10.25302/12.2020.AD.131008215

16. Zheng J, Wittouck S, Salvetti E, et al. A taxonomic note on the genus Lactobacillus: Description of 23 novel genera, emended description of the genus Lactobacillus Beijerinck 1901, and union of Lactobacillaceae and Leuconostocaceae. Int J Syst Evol Microbiol. 2020;70(4):2782–2858. doi:10.1099/ijsem.0.004107

17. Tractenberg RE, Frost JK, Yumoto F, Rounds AK, Ljungberg IH, Groah SL. Validity of the Urinary Symptom Questionnaires for people with neurogenic bladder (USQNB) who void or use indwelling catheters. Spinal Cord. 2021;59(9):948–958. doi:10.1038/s41393-021-00666-w

18. Tractenberg RE, Frost JK, Yumoto F, Rounds AK, Ljungberg IH, Groah SL. Reliability of the Urinary Symptom Questionnaires for people with neurogenic bladder (USQNB) who void or use indwelling catheters. Spinal Cord. 2021;59(9):939–947. doi:10.1038/s41393-021-00665-x

19. Hooton TM, Bradley SF, Cardenas DD, et al. Diagnosis, Prevention, and Treatment of Catheter-Associated Urinary Tract Infection in Adults: 2009 International Clinical Practice Guidelines from the Infectious Diseases Society of America. Clin Infect Dis. 2010;50(5):625–663. doi:10.1086/650482

20. Fouts DE, Pieper R, Szpakowski S, et al. Integrated next-generation sequencing of 16S rDNA and metaproteomics differentiate the healthy urine microbiome from asymptomatic bacteriuria in neuropathic bladder associated with spinal cord injury. J Transl Med. 2012;10(1):1. doi:10.1186/1479-5876-10-174

21. Groah SL, Pérez-Losada M, Caldovic L, et al. Redefining Healthy Urine: A Cross-Sectional Exploratory Metagenomic Study of People With and Without Bladder Dysfunction. J Urol. 2016;196(2):2. doi:10.1016/j.juro.2016.01.088

22. Groah SL, Rounds AK, Ljungberg IH, Sprague BM, Frost JK, Tractenberg RE. Intravesical Lactobacillus rhamnosus GG is safe and well tolerated in adults and children with neurogenic lower urinary tract dysfunction: first-in-human trial. Ther Adv Urol. 2019;11:1756287219875594. doi:10.1177/1756287219875594

23. Forster CS, Hsieh MH, Pérez-Losada M, et al. A single intravesical instillation of *Lactobacillus rhamnosus* GG is safe in children and adults with neuropathic bladder: A phase Ia clinical trial. J Spinal Cord Med. 2021;44(1):62–69. doi:10.1080/10790268.2019.1616456

24. Tractenberg RE, Groah SL, Frost JK, et al. Effects of Intravesical Lactobacillus Rhamnosus GG on Urinary Symptom Burden in People with Neurogenic Lower Urinary Tract Dysfunction. PM R. Published online August 14, 2020. doi:10.1002/pmrj.12470

25. Tractenberg RE, Groah SL, Rounds AK, Ljungberg IH, Schladen MM. Preliminary validation of a Urinary Symptom Questionnaire for individuals with Neuropathic Bladder using Intermittent Catheterization (USQNB-IC): A patient-centered patient reported outcome. PLOS ONE. 2018;13(7):7. doi:10.1371/journal.pone.0197568

26. Parikh CR, Butrymowicz I, Yu A, et al. Urine stability studies for novel biomarkers of acute kidney injury. Am J Kidney Dis Off J Natl Kidney Found. 2014;63(4):4. doi:10.1053/j.ajkd.2013.09.013

27. Schuh MP, Nehus E, Ma Q, et al. Long-term Stability of Urinary Biomarkers of Acute Kidney Injury in Children. Am J Kidney Dis Off J Natl Kidney Found. 2016;67(1):1. doi:10.1053/j.ajkd.2015.04.040

28. Reid G, Bruce AW. Probiotics to prevent urinary tract infections: the rationale and evidence. World J Urol. 2006;24(1):28-32. doi:10.1007/s00345-005-0043-1

29. Cadieux PA, Burton J, Devillard E, Reid G. Lactobacillus by-products inhibit the growth and virulence of uropathogenic Escherichia coli. J Physiol Pharmacol Off J Pol Physiol Soc. 2009;60 Suppl 6:13–18.

30. Abdul-Rahim O, Wu Q, Price TK, et al. Phenyl-lactic acid is an active ingredientin bactericidal supernatants of *Lactobacillus crispatus*. J Bacteriol. Published online July 19, 2021. doi:10.1128/JB.00360-21

31. Karlsson M, Scherbak N, Reid G, Jass J. Lactobacillus rhamnosus GR-1 enhances NF-kappaB activation in Escherichia coli-stimulated urinary bladder cells through TLR4. BMC Microbiol. 2012;12(1):15. doi:10.1186/1471-2180-12-15

32. Anukam KC, Hayes K, Summers K, Reid G. Probiotic *Lactobacillus rhamnosus* GR-1 and *Lactobacillus reuteri* RC-14 May Help Downregulate TNF-Alpha, IL-6, IL-8, IL-10 and IL-12 (p70) in the Neurogenic Bladder of Spinal Cord Injured Patient with Urinary Tract Infections: A Two-Case Study. Adv Urol. 2009;2009:1–5. doi:10.1155/2009/680363

33. Groah SL, Rounds AK, Pérez-Losada M. Intravesical Lactobacillus rhamnosus GG Alters Urobiome Composition and Diversity Among People With Neurogenic Lower Urinary Tract Dysfunction. Top Spinal Cord Inj Rehabil. 2023;29(3):44–57. doi:10.46292/sci23-00004

34. Groah S, Tractenberg RE, Frost JK, Rounds A, Ljungberg I. Independence of Urinary Symptoms and Urinary Dipstick Results in Voiders With Neurogenic Bladder. Top Spinal Cord Inj Rehabil. 2022;28(2):2. doi:10.46292/sci21-00006

35. Rounds AK, Tractenberg RE, Groah SL, et al. Urinary Symptoms Are Unrelated to Leukocyte Esterase and Nitrite Among Indwelling Catheter Users. Top Spinal Cord Inj Rehabil. 2023;29(1):1. doi:10.46292/sci22-00095

36. Tractenberg RE, Groah SL, Newcomb E, Riegner C, Forster C. Normal Variability of Urinary Markers in Individuals with Spinal Cord Injury and Disease Using Intermittent Catheterization (in prep). Preprint posted online February 4, 2026. doi:10.64898/2026.02.02.26345397

37. Cohen CR, Wierzbicki MR, French AL, et al. Randomized Trial of Lactin-V to Prevent Recurrence of Bacterial Vaginosis. N Engl J Med. 2020;382(20):1906–1915. doi:10.1056/NEJMoa1915254

